# Closing the accessibility gap to mental health treatment with a conversational AI-enabled self-referral tool

**DOI:** 10.1101/2023.04.29.23289204

**Authors:** Johanna Habicht, Sruthi Viswanathan, Ben Carrington, Tobias Hauser, Ross Harper, Max Rollwage

## Abstract

Accessing mental health care can be challenging, and minority groups often face additional barriers. This study investigates whether digital tools can enhance equality of access to mental health treatment. We evaluated a novel AI-enabled self-referral tool (a chatbot) designed to make entry to mental health treatment more accessible in a real-world setting.

In a multi-site observational study, data were collected from 129,400 patients who referred to 28 separate NHS Talking Therapies services across England. Our results indicate that the tool led to a 15% increase in total referrals, which was significantly larger than the 6% baseline increase observed in matched services using traditional self-referral methods during the same time period. Importantly, the tool was particularly effective for minority groups, which included non-binary (235% increase), bisexual (30% increase), and ethnic minority individuals (31% increase). This paints a promising picture for the use of AI chatbots in mental healthcare and suggests they may be especially beneficial for demographic groups that experience barriers to accessing treatment in the traditional care systems.

To better understand the reasons for this disproportional benefit for minority groups, we used thematic analysis and Natural Language Processing (NLP) models to evaluate qualitative feedback from 42,332 individuals who referred through the AI-enabled tool. We found that the tool’s human-free nature and its ability to improve the perceived need for treatment were the main drivers for improved diversity.

These findings suggest that AI-enabled chatbots have the potential to increase accessibility to mental health services for all, and to alleviate barriers faced by disadvantaged populations. The results have important implications for healthcare policy, clinical practice, and technology development.

## 1 Introduction

Mental health is a global health priority, with the World Health Organization (WHO) recognising that mental health conditions are one of the leading causes of disability and disease burden worldwide (World Health Organisation, 2022). The most prevalent mental health disorders, such as anxiety disorders and depression, were estimated to affect 29% of the world’s population in their lifetime before the COVID-19 pandemic [Steel et al., 2014]. The pandemic further exacerbated the mental health crisis and increased the need for more support [Thome et al., 2021, Ornell et al., 2021, Loosen et al., 2021, Santomauro et al., 2021]. Besides the personal implications of mental health problems, anxiety disorders and depression alone lead to decreased productivity, causing the global economy to lose US $1 trillion annually [Health, 2020]. By 2030, poor mental health is projected to cost $6 trillion, underlining the urgent need to find solutions to address this global crisis [Health, 2020].

Mental health support can reduce the burden and has been found to be highly effective for many mental disorders. However, two main challenges preclude access to mental health support for many. Firstly, the supply of mental healthcare is not matching the demand as mental health services are widely underfunded and understaffed [World Health Organization, 2021b, Mahase, 2020]. Secondly, not every individual experiencing mental health problems seeks help, or they delay seeking it for years [Wang et al., 2007, Department of Health and Social Care, 2023], resulting in unmet needs and inadequate support at the right time. This can be due to barriers, such as a lack of perceived need for treatment, negative attitudes and stigma around mental health, as well as structural barriers [Andrade et al., 2014, Thornicroft et al., 2022]. These barriers can be even stronger for individuals among minority and disadvantaged backgrounds that may be stigmatised, such as individuals with ethnic or sexuality minority status [Clement et al., 2015, Thornicroft et al., 2022]. Digital technologies and Artificial Intelligence (AI) have been proposed as potential solutions to these challenges [Rudd and Beidas, 2020, Taylor et al., 2020, Kola et al., 2021]. While such technology has been shown to reduce the workload of the mental health staff and make services more efficient [Jayaraajan et al., 2022, Koutsouleris et al., 2022, Rollwage et al., 2022], less is known about the marginal impact of AI technologies in supporting individuals of differing demographics seeking help. Excitingly, digital solutions might be especially well-positioned to help individuals who are seeking support to overcome barriers, for example by offering more flexibility and reducing stigma [Rodríguez-Rivas et al., 2022].

The first step of many mental health care pathways is for individuals to seek help and be referred to the appropriate health care service. Ensuring easy and inclusive access at this initial step is crucial, regardless of an individual’s socioeconomic status, ethnicity, gender, or other such factors. This underpins a fair and equitable healthcare system. Unfortunately, evidence suggests that individuals from minority groups often face higher barriers and stigma in accessing care [Corrigan et al., 2014]. Referral to the appropriate healthcare service is a pivotal point in the treatment pathway, as failure to access the right support at the right time can lead to a worsening of symptoms, comorbidities, and adverse outcomes, including hospitalization or suicide [Williams et al., 2008, Reichert and Jacobs, 2018, De Girolamo et al., 2012]. Notably, a significant proportion of mental health problems remain undiagnosed and untreated, with estimates suggesting that as many as two-thirds of individuals with depression do not receive the necessary care [Moitra et al., 2022]. Therefore, making help-seeking more accessible is critical to providing timely and appropriate care to individuals in need, promoting better outcomes, and enhancing overall well-being.

The referral process for mental health care services varies between countries and services, and can involve self-referral or referral by a medical professional such as a general practitioner [Volpe et al., 2015]. The more efficient the referral process, the quicker support can be accessed, reducing the burden for the individual and the wider society. In the United Kingdom, the NHS Talking Therapies for Anxiety and Depression program - formerly Improving Access to Psychological Therapies (IAPT) - predominantly relies on self-referrals [Brown et al., 2010, The National Collaborating Centre for Mental Health, 2021]. Of the 1,740,652 patients referred to NHS Talking Therapies in 2021, 72% were self-referrals (NHS Digital). However, existing solutions may not be optimised or digitally streamlined, leading to lower completion rates and limited access to services [NHS Digital, 2019]. Thus, more optimal solutions are needed, as well as research into the evidence for these solutions.

To optimise this self-referral process, digital technologies have been suggested as a solution, such as AI chatbot solutions that have gained increased attention in healthcare [Tudor Car et al., 2020]. There is growing evidence that technology can improve help-seeking behaviour [Johnson et al., 2022] and that patients prefer interacting with chatbots for self-reported measures [Te Pas et al., 2020] and discussing sensitive topics [Bhakta et al., 2014]. This suggests that digital technologies can remove some of the barriers to accessing mental health services, and thus increase accessibility.

To this end, we developed a novel AI-enabled chatbot solution for self-referrals, *Limbic Access*, which can optimise the standard referral process by autonomously gathering patient information to inform suitability for the service and initial presenting problem. We hypothesised that this AI-augmented self-referral could lower the threshold for accessing mental health services for three main reasons. Firstly, it can eliminate access barriers by providing a more convenient and flexible option than existing self-referral methods, such as via phone calls or static, daunting, and difficult-to-find webforms hosted on the service website. Additionally, the AI-enabled self-referral can be completed outside of service hours and with greater privacy. Secondly, a chatbot can guide patients proactively through the referral process, improving completion rates. Finally, chatbots have the potential to reduce the negative attitudes and stigma surrounding mental health and help-seeking. Therefore, we anticipated an increase in total referrals to services using the novel AI-enabled self-referral tool, as well as an increase in referrals from minority groups who face more barriers to access.

The use of AI chatbot solutions in healthcare is still an emerging area of research [Wilson and Marasoiu, 2022], with limited studies conducted in clinical settings. Therefore, our study aimed to assess the effectiveness of an AI-enabled self-referral tool for increasing accessibility to mental health services in a clinical context. To achieve this, we conducted an observational study using real-world data from the UK — a suitable environment for testing the self-referral tool due to its standardized referral process and a high percentage of self-referrals to NHS Talking Therapies.

Our results show that the AI-enabled self-referral tool, implemented in 14 services, had a positive impact on the number and diversity of referrals in a real-world setting. To further investigate the factors behind the improvement in diversity of access, we conducted thematic analysis on the qualitative feedback of 42,332 patients who used the tool, using Natural Language Processing (NLP) models. Our analysis showed that the tool’s ease of use and convenience were the primary drivers of the increase in referrals. Additionally, for minority groups, we identified two significant drivers: the tool’s human-free nature, which allowed patients to seek help without fear of being judged or discriminated against, and its ability to increase patients’ awareness of their treatment needs. These findings suggest that the tool may be able to alleviate the stigma experienced by minority groups when seeking mental healthcare

## 2 Method

Here we studied the effects of an AI-enabled self-referral tool, *Limbic Access*, on the number and diversity of referrals to the NHS Talking Therapies for Anxiety and Depression (previously Improving Access to Psychological Therapies (IAPT) programme) in the United Kingdom. This tool has been developed with NHS Talking Therapies services and has been implemented as part of the routine mental healthcare in several NHS Talking Therapies services.

### 2.1 AI-enabled self-referral tool

The AI-enabled self-referral tool has been integrated into the service’s website (see Figure 1B) as a chatbot which is visible to any individual who visits the website. This reduces the barrier to seeking help as users do not need to manually search for how to get in contact with the service. The self-referral tool collects the necessary information for a referral required by the NHS Talking Therapies services (e.g. eligibility criteria, contact details, and demographic information). Additionally, the chatbot collects clinical information about the patient’s presenting symptoms, using questionnaires such as the Patient Health Questionnaire-9 (PHQ-9) [Kroenke et al., 2001], Generalised Anxiety Disorder Assessment (GAD-7) [Spitzer et al., 2006], the Work and Social Adjustment Scale (WSAS) [Mundt et al., 2002], and additional screening questions. These routine questionnaires and screening questions have not been typically collected at the point of referral in NHS Talking Therapies, but rather during the first assessment on a phone call. Frontloading the questionnaires during the referral process can reduce administrative burden, as well as make it easier for individuals to fill out these questionnaires in their own time and without having to speak to a person. Once the individual has completed the referral information using the chatbot, all the data is attached to the referral record within the NHS Talking Therapies services’ electronic health record to support the clinician in providing a high-quality and high-efficiency clinical assessment.

**Figure 1:**
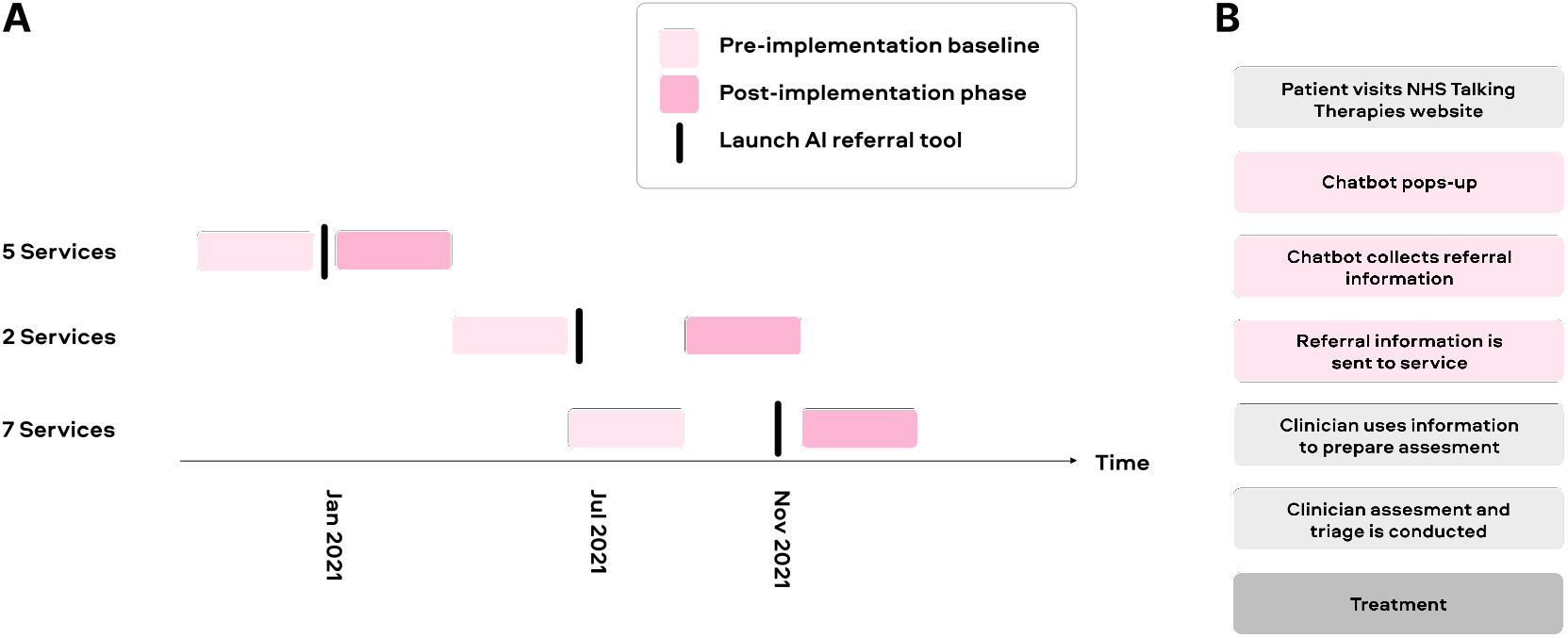
A) Study design for a real-world retrospective observational study with multiple sites, showing a three-month pre-implementation and three-month post-implementation period for different services. B) Referral workflow, showing the patient’s journey from visiting the NHS Talking Therapies service’s website to entering treatment.

### 2.2 Study design

This is a multi-site real-world retrospective observational study to investigate the impact of the AI-enabled self-referral tool on the number and diversity of referrals. The data used in this study are recorded in routine practice by all NHS Talking Therapies services and are reported to the public monthly, quarterly, and annually by NHS Digital (NHS Digital). The publicly available data undergoes quality checks and is a comprehensive dataset that facilitates high-quality analysis. The data used for the total number of referrals are taken from the annual reports and the demographic data is taken from the quarterly reports.

The analysis was conducted using data from ∼129,400 patients who referred to 28 different NHS Talking Therapies services across England: 14 of these services implemented the AI-enabled self-referral tool and 14 of them did not while matching for the same timeframe. To understand the effects of the novel self-referral tool on the number and diversity of referrals, we investigated the time period before (pre-AI-enabled self-referral tool) and after the launch of the self-referral tool (post-AI-enabled self-referral tool) in the services. The timeframes for the preand post-periods were chosen as the closest quarter (3 months) to when the service launched the AI-enabled self-referral tool (see Figure 1A). Different services launched the self-referral tool at different times, allowing for a staggered study design to account for the fluctuation of referrals throughout the year due to seasonal and other factors. To ensure that the effects observed were not confounded by other factors, we matched the services that implemented the AI-enabled self-referral tool to other similar services as commonly done in similar clinical and public health research studies [Rose and Van der Laan, 2009]. We only included control services with a webform as one of their ways of referral because this was the closest referral option to the AI-enabled self-referral tool investigated, thus representing an optimal control group. For each NHS Talking Therapies service using the AI-enabled self-referral tool, we calculated the total number of referrals in the pre- AI self-referral tool period. Then we calculated the total number of referrals for all other NHS Talking Therapies services during the same period of time. For example, when the tool was launched in July 2021, we calculated the number of total referrals for the second quarter (April to June) of 2021 for all services and used these metrics for matching the most similar control services. In order to match services, we calculated the Euclidean distance based on these two (normalized) features and took the service with the lowest distance to the NHS Talking Therapies service using the AI-enabled self-referral tool, where a low distance implies a high degree of similarity.

### 2.3 Analysis

#### 2.3.1 Total number of referrals

To investigate whether the AI-enabled self-referral tool increased the number of referrals, we calculated the total number of referrals for each service in the pre- and post-implementation of the tool. To analyse the data between the pre- and post-implementation periods, we summed the total number of referrals for all services. To compare the change in the total number of referrals between services that implemented the AI-enabled self-referral tool and those that did not, we used a Chi-squared test.

To ensure that the total number of referrals did not increase due to other forms of referrals other than self-referrals (e.g. GP referrals) increasing in the time period, we repeated the analysis for only self-referrals in the same services at the same time period (Supplementary Figure 1).

#### 2.3.2 Demographic data

To investigate whether the AI-enabled self-referral tool improved the diversity of referrals, we investigated the number of referrals for certain sociodemographic groups, focusing on gender identity, sexual orientation and ethnic groups. Specifically, we investigated the number of referrals for females, males, and non-binary individuals for gender identity; heterosexual, bisexual, and gay/lesbian individuals for sexual orientation; and Asian or Asian British, Black or Black British, Mixed, Other Ethnic groups, and White individuals for ethnicity. We calculated the total number of referrals from a certain sociodemographic group for the pre- and post-implementation periods and the percentage change for each group.

To further assess whether increased referrals from minority groups exceeded those of the most common groups, we constructed a logistic regression predicting on a single, inferred, patient level whether the patient referred pre-or post-implementation of the tool (0 = pre-implementation, 1 = post-implementation). As a predictor variable, we used the sociodemographic group of the individual, assigning the most common group (e.g. females for gender, heterosexuals for sexual orientation and white for an ethnic group) as the reference. We report the odds ratio, calculated as the exponential of the regression coefficient, along with confidence intervals.

#### 2.3.3 NLP topic classification

To understand the mechanisms behind the increased access, we used thematic analysis powered NLP methods to analyse qualitative feedback given by patients who used the AI-enabled self-referral tool. NLP techniques, such as text classification, have gained popularity in healthcare settings for analysing free-text patient feedback to generate insight about patients’ experience [Khanbhai et al., 2021]. In this study, we employed a supervised NLP classification model to categorise qualitative feedback provided by individuals upon completing their referrals via the AI-enabled self-referral tool to better understand the reasons for the increased number of referrals, especially from minority groups. From September 2021 to March 2023, a total of 157,416 patients used this novel self-referral approach, with 29% of these patients offering free-text feedback on their experience (N = 46,166). To ensure our analysis only included meaningful insights, we excluded any responses containing fewer than 10 characters, resulting in a final sample of 42,332 feedback entries. We employed a NLP classification model to categorize each feedback entry to a theme.

We first performed reflexive thematic analysis on feedback entries [Braun and Clarke, 2019], with one researcher open-coding a sample of 4,000 entries. Initial codes were discussed with the larger group of researchers, and a consensus was reached on the resulting themes (listed in the table below) after two meetings. Our team manually identified nine distinct themes, consisting of four positive themes and two neutral and three negative themes. We then selected entries under each theme to train a model on a subset of feedback entries (657 entries), ensuring that each theme had at least 50 representative entries. This subset was then utilised as the training and testing dataset for the NLP model.

| Theme | Explanation |
| --- | --- |
| Convenient | Individuals mentioned that the referral process was easy and/or fast |
| Provided hope | Individuals mentioned that the referral was a hopeful first step towards getting support |
| Self-realisation | Individuals mentioned that the referral provided a self-realisation of their current situation, such as a realisation of suffering from a mental health condition or a need for treatment |
| Human-free | Individuals mentioned the human-free nature of the tool and removing the anxiety of talking to humans |
| Needed specific support | Individuals mentioned that they needed to talk about specific illnesses or other mental disorders |
| Other neutral feedback | Individuals mentioned any other neutral feedback |
| Expected support sooner | Individuals mentioned that they expected to start therapy faster |
| Wanted urgent support | Individuals mentioned that they felt that they needed immediate attention. |
| Other negative feedback | Individuals mentioned any other negative feedback |

To prepare the feedback data for use with our model, we first used a sentence-BERT (SBERT) sentence transformer to transform each entry into numerical representations. We then employed Principal Component Analysis (PCA) to reduce dimensions. We used a multi-class logistic regression model and evaluated its accuracy through a 100-fold cross-validation approach. Our model achieved a micro-average F1 score of 0.64. This suggests that the model demonstrated a moderate level of performance in classifying patient feedback entries into their assigned themes.

To classify the 42,332 feedback entries into the nine themes, we employed the above-mentioned model. To determine whether the proportions of individuals mentioning specific themes differed between minority and majority groups, we used a Chi-squared test. To account for multiple comparisons, we used Bonferroni correction.

## 3 Results

### 3.1 Increase in the number and diversity of referrals

The implementation of the AI-enabled self-referral tool resulted in a 15% increase in the total number of referrals compared to a similar pre-implementation time period (Figure 2). In contrast, matched NHS Talking Therapies services with a similar number of total referrals in the pre-implementation period that used other self-referral methods, such as web forms, saw only a 6% increase in referrals in the same time period. The increase in total referral numbers for services using the AI-enabled self-referral tool was significantly higher than the increase for the matched control services (*χ*^2^ (1) = 86.3, *p <* 0.0001). This suggests that the AI-enabled self-referral tool facilitates an increase in referrals compared to the national average in matched NHS Talking Therapy services during the same time period.

**Figure 2:**
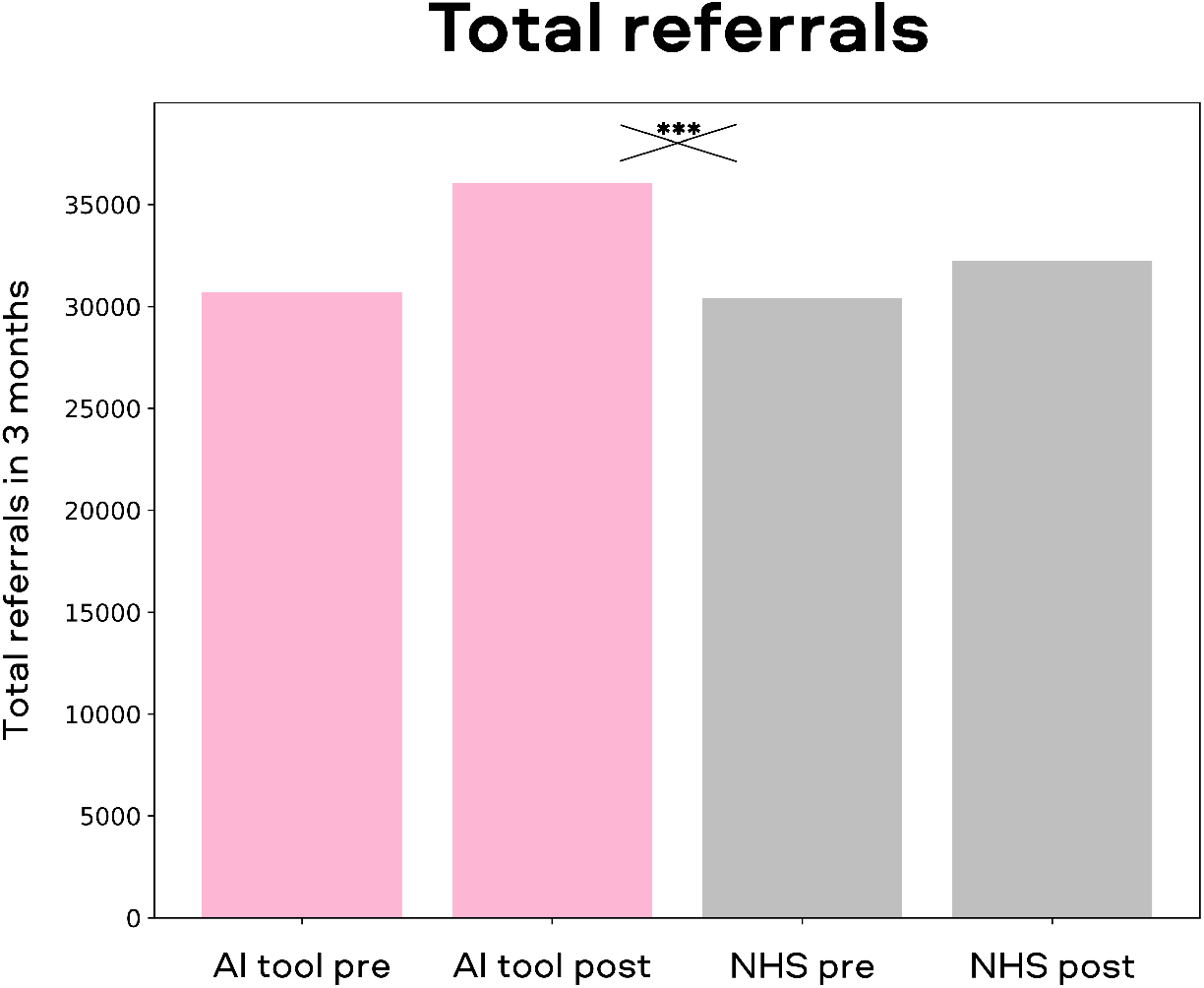
The total number of referrals pre- and post-implementation of the AI-enabled self-referral tool for services that implemented the novel AI tool (pink) and other similar services that did not implement the tool (grey). There was a 15% increase in total referrals for services that used the AI tool, whereas other services had a 6% increase in the same time period.The increase in referrals for services using the self-referral tool was significantly higher than the increase for other similar services.\*\*\**p <* 0.001

The results suggest that the AI-enabled self-referral tool has a generally positive impact on increasing access to mental health treatment for individuals. However, some disadvantaged sociodemographic groups may encounter more barriers when trying to access treatment. To explore this further, we investigated whether the tool’s effect on increasing referrals holds true across multiple demographic groups or if it disproportionately benefits certain groups in accessing mental health care. We calculated the percentage change in the number of referrals from preto post-implementation of the AI-enabled self-referral tool, separately for all sociodemographic groups (see Supplementary Table 1 for the total number of referrals during pre- and post-implementation) and compared minority groups to the most common groups.

First, we focused on the gender identity of individuals who referred to the services. We found that referrals from individuals who identified as non-binary increased by 235%, whereas referrals from individuals who identified as female or male increased by 18% and 16%, respectively (Figure 3A). The increase in referrals from people who identify as non-binary was higher than females (odds-ratio = 2.83, CI = [2.264,3.545], *p <* 0.0001) and males (odds-ratio = 2.90, CI = [2.314,3.629], *p <* 0.0001), suggesting that the AI-enabled self-referral tool especially facilitates more access for people from gender minority groups.

**Figure 3:**
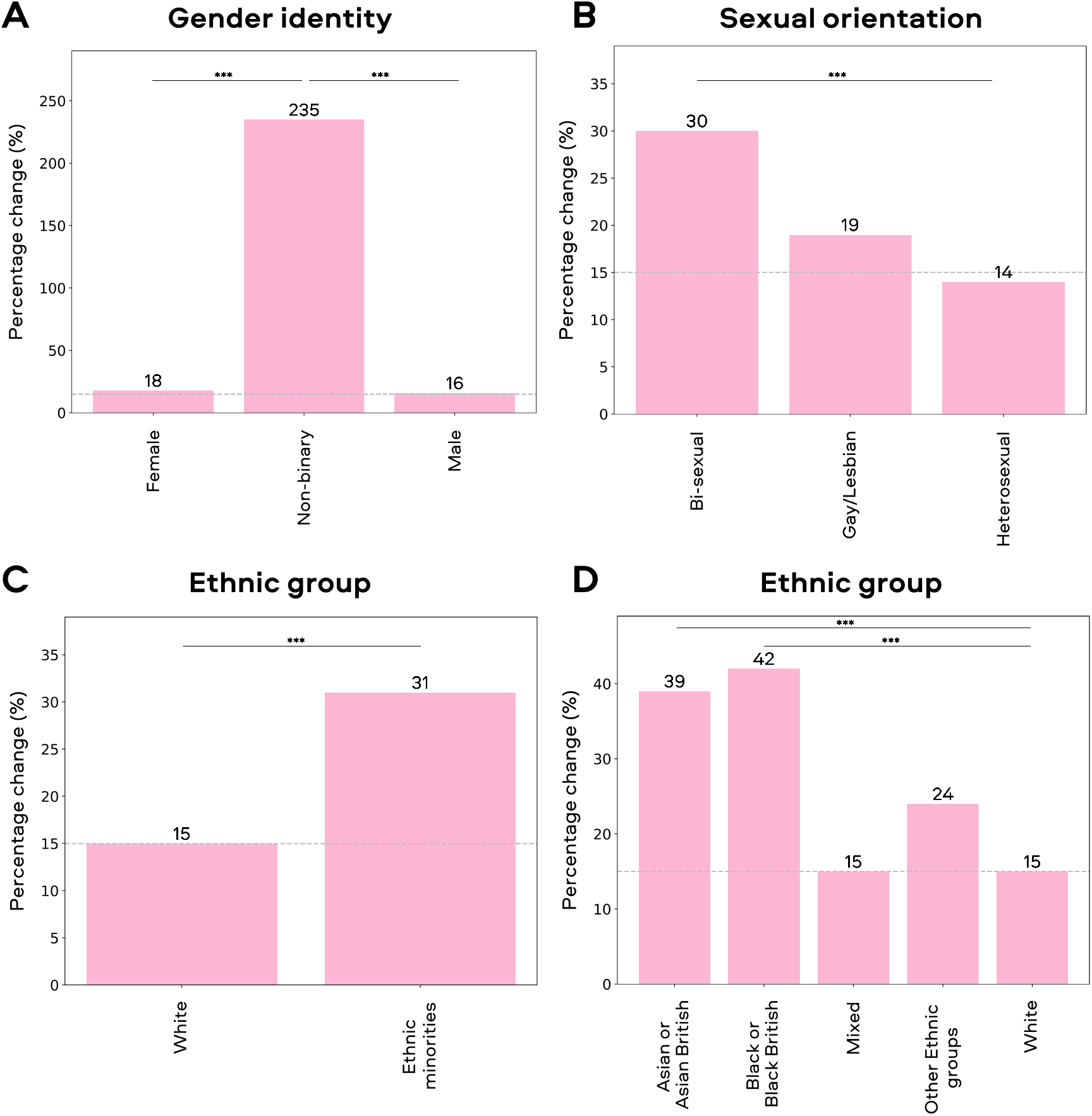
Percentage change for different sociodemographic groups for services that implemented the AI-enabled self-referral tool. The dotted line on each plot shows the overall percentage change in the total referral numbers (15%). A) A positive percentage change in referrals was observed for all genders, with the highest percentage increase in referrals for individuals who identify as non-binary. B) A positive percentage change in referrals was observed for people with all sexual orientations, with the highest percentage increase in referrals for individuals who are bi-sexual. C) A positive percentage change in referrals was observed for all ethnic groups, with the highest percentage increase in referrals for ethnic minorities. D) There was a higher positive change for Asian or Asian British and Black or Black British compared to the White group. \*\*\**p <* 0.001

Next, we investigated referrals based on individuals’ sexuality. We found that referrals by people whose sexual orientation was bi-sexual increased by 30%, referrals by people whose sexual orientation is gay/lesbian increased by 19%, whereas referrals from heterosexuals increased by 14% (Figure 3B). The increase of people whose sexual orientation is bi-sexual was higher than for heterosexual patients (odds-ratio = 1.14, CI= [1.074,1.219], *p* =*<* 0.0001), suggesting that the tool facilitates more access for people who are not heterosexual. However, the increase in individuals identifying as homosexual did not significantly differ from the increase in referrals for heterosexual individuals (odds-ratio = 1.05, CI= [0.922,1.189], *p* =*<* 0.216).

Lastly, we evaluated the effects of the tool on referral rates for different ethnic groups. We identified a 31% increase in referrals for ethnic minorities, whereas the increase for the White group was 15% (Figure 3C), suggesting that different ethnic groups were more represented after the implementation of the AI-enabled self-referral tool. The increase in ethnic minorities was higher than for White individuals (odds-ratio = 1.14, CI= [1.081,1.202], *p <* 0.0001), suggesting that the tool facilitates more access for people from ethnic minorities. To investigate these results further, we analysed ethnic groups in a more fine-grained manner. We particularly saw an increase for Asian and Asian British groups (odds-ratio =1.21, CI = [1.116,1.318], *p <* 0.0001) and Black and Black British groups (odds-ratio = 1.24, CI = [1.108,1.378], *p <* 0.0001) compared to White individuals (Figure 3D). We did not see any differences in the number of referrals for individuals from other ethnic groups (odds-ratio =1.08, CI = [0.913,1.272], *p* = 0.377) and Mixed ethnic group (odds-ratio =1.01, CI = [0.917,1.107], *p* = 0.877) compared to White individuals.

In conclusion, these results suggest that the AI-enabled self-referral tool facilitates a general increase in referrals, as well as improves the diversity of referrals. An increase in referrals can potentially lead to longer wait times for clinical assessment and may reduce the number of people who access these assessments. To address this concern, we compared the percentage of people who were assessed in the time periods between the services that used the AI-enabled self-referral tool and services not using the tool (Supplementary Figure 3). A two-way ANOVA revealed no significant interaction between the time period and tool usage (F(1, 13) = 0.09, *p* = 0.764). Therefore, the AI-enabled self-referral tool did not negatively impact the number of people who accessed assessments.

### 3.2 Mechanism for the increase in referrals

To investigate the underlying mechanism behind the increased number and diversity of referrals through the AI-enabled self-referral tool, we analysed qualitative free-text feedback provided by 42,332 individuals upon completion of their referral. Our goal was to identify whether specific topics, such as specific advantages or disadvantages of the AI-enabled self-referral tool, were mentioned disproportionately by certain demographic groups. We used NLP topic classification to classify each feedback entry to a predefined theme (see Methods for details). We analysed feedback from individuals belonging to both minority and majority groups based on their ethnic background, sexual orientation, and gender identity. We compared minority groups where we observed differences in the number of referrals bisexual individuals, non-binary and other gender minority individuals, and Asian, Black and other ethnic group individuals (Figure 4). We included the analysis of all minority groups in the Supplementary material (Supplementary Figure 2).

**Figure 4:**
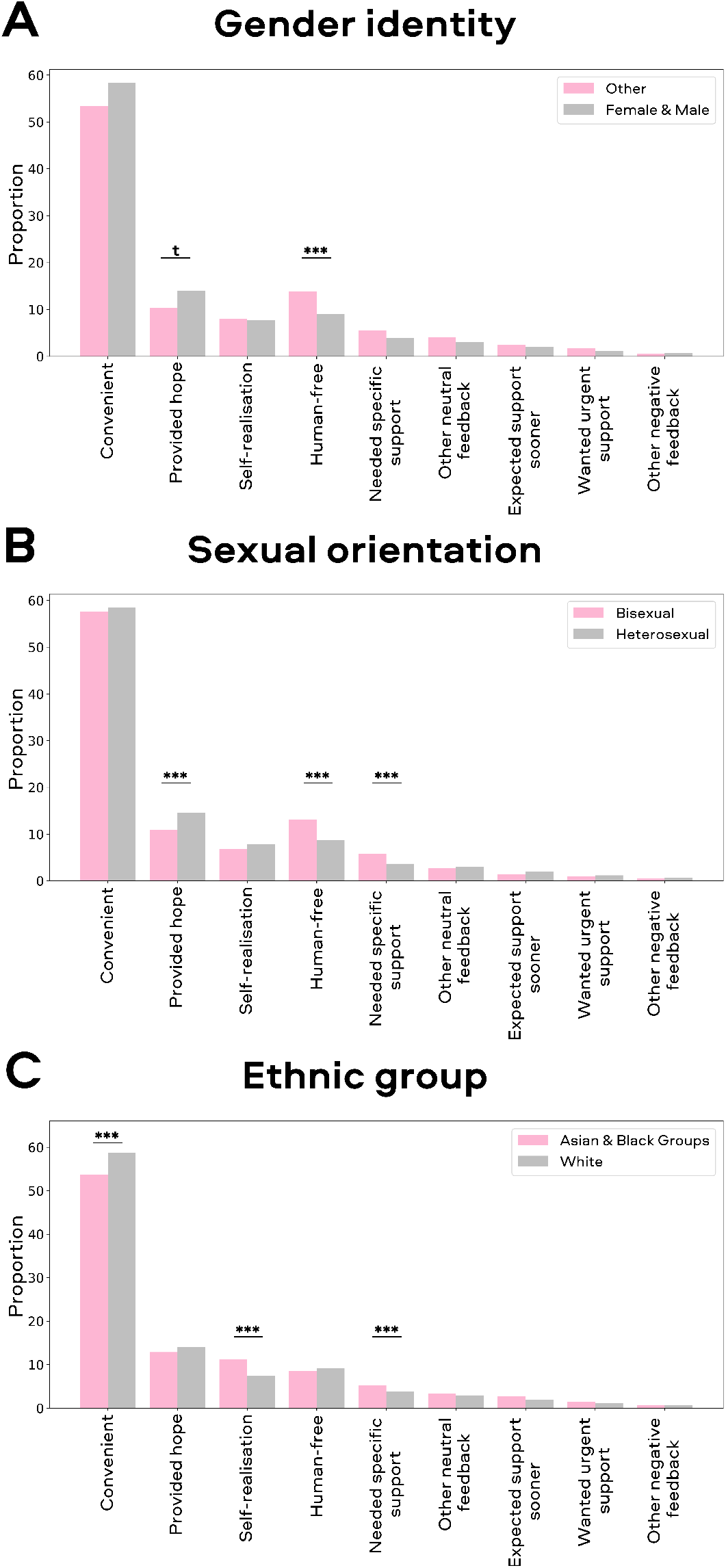
Breakdown of feedback themes from minority (pink bars) and majority (grey bars) groups. A) Individuals from gender minority groups mentioned the tool’s human-free nature more compared to the majority groups. B) Bisexual individuals mentioned the tool’s human-free nature more, as well as needing more specific support compared to the majority groups. C) Individuals from Asian and Black ethnic groups mentioned more the theme around self-realisation as well as needing more specific support compared to White individuals. \*\*\**p <* 0.001; ** *p <* 0.01; t *p <* 0.1

First, it is noteworthy that there are some generally overarching patterns across all demographic groups. Overall, 89% of the free text feedback was classified as positive, 7% neutral and 4% negative. In general, patients mentioned the convenient nature of the chatbot as the major advantage, emphasizing the effectiveness of this novel self-referral method. This was followed by the hope of having taken the first step to access support, the absence of human involvement reducing stigma and judgment and increased self-realization that they might suffer from a mental health condition and need support caused by the self-referral process. While these general patterns largely hold across all demographic groups, there were significant differences in the frequency with which these advantages were mentioned, indicating that different groups might experience different advantages which might contribute to the increased accessibility for these groups. The following sections highlight the significant findings, while additional results can be found in Supplementary Table 2-4.

We found that individuals from gender minority groups mentioned the absence of human involvement in the tool more frequently than females and males (*χ*^2^ (1) = 22.7, *p <* 0.0001, Figure 4A). This suggests that the AI-enabled self-referral tool resolves an important barrier for this group by being human-free and indicates the mechanism for why we observed increased referrals from this minority group might be a reduction in stigma and judgment during this AI-enabled self-referral process. We observed a marginally significant reduction in the theme related to the self-referral providing hope for the gender minority groups (*χ*^2^ (1) = 8.84, *p* = 0.079), which may suggest that the absence of human involvement in the tool is a more important factor for this group than the sense of hope it provides.

Similarly, we found that individuals who identify as bisexual mentioned the human-free nature of the tool more frequently than heterosexual individuals (*χ*^2^ (1) = 62.4, *p <* 0.0001, Figure 4B) indicating that the human-free aspect is important for them. In addition, we saw a similar reduction in the theme about hope for bisexual individuals,(*χ*^2^ (1) = 27.8, *p <* 0.0001) indicating that the human-free aspect is more important for them compared to the feeling of hope provided by the tool.

When investigating ethnic minorities, we found that individuals from Asian and Black ethnic groups mentioned the self-realisation about the need for treatment theme more than White individuals (*χ*^2^ (1) = 46.6, *p <* 0.0001, Figure 4C), which reflects that during the self-referral process, there was an increase in the individual’s awareness that they might suffer from a mental health condition and the need for treatment. This may reflect how the tool can reduce the stigma faced by ethnic minority groups in seeking mental health support. In addition, we found that Asian and Black ethnic minority groups mentioned the convenient nature of the tool less than White individuals (*χ*^2^ (1) = 24.2, *p <* 0.0001). Finally, our analysis showed that individuals from Asian and Black ethnic groups and bisexual individuals provided more neutral feedback compared to majority groups, especially around the theme of needing specific support (sexuality: *χ*^2^ (1) = 30.8, *p <* 0.0001; ethnicity: *χ*^2^ (1) = 14.6, *p* = 0.0036). This could be attributed to the higher barriers faced by these individuals in seeking mental health support in general, as well as their specific problems related to their minority status, such as bisexual individuals seeking help for specific problems related to their sexuality.

In conclusion, our findings suggest that minority groups may face different barriers and value different aspects when seeking mental health treatment than majority groups. Some of these barriers might be especially well addressed by an AI-enabled self-referral tool’s human-free aspect and the reduced stigma. Such solutions may remove these barriers and may be valued more by these minority groups, leading to a higher number of referrals.

## 4 Discussion

In this study, we investigated the effects of an AI-enabled self-referral tool on the number and diversity of referrals in the NHS Talking Therapies services in the United Kingdom. Notably, this is the first large-scale study using real-world data to investigate the potential impact of an AI-enabled tool on the quality of access to mental health care. We found that NHS Talking Therapies services that implemented the AI-enabled self-referral tool showed a 9 percentage point stronger increase in the number of referrals after the launch of this tool compared to matched NHS Talking Therapies services that used other forms of referral in the same time period. Specifically, services that used the AI-enabled self-referral tool showed a 15% increase in referrals, whereas the matched services that did not use the tool showed a 6% increase. This baseline increase is likely to reflect the national initiative to boost access across all NHS Talking Therapies services. At the time of writing, this self-referral tool has facilitated the referral of ∼157,000 patients, which means that ∼14,000 additional patients have found their way into treatment owing to this technical solution. Importantly, the increase in referrals did not negatively impact the number of individuals who accessed clinical assessments, which suggests that the tool did not exacerbate wait times for treatment. In addition, a separate study involving the same services demonstrated that this AI-enabled self-referral tool has further downstream implications and improves recovery rates [Rollwage et al., 2022]

An important insight from our study is that the services using the AI-enabled self-referral tool saw a proportionally higher increase in the number of individuals from minority groups, such as individuals who identify as non-binary, bisexual, and ethnic minorities. For individuals from ethnic minority backgrounds, we especially saw an increase for Asian or Asian British groups and Black or Black British groups. These groups have been previously underrepresented in mental healthcare treatment, indicating that this tool enabled a previously underserved need.

Findings from previous research suggest that digital technologies may facilitate help-seeking for mental health [Johnson et al., 2022] and make it easier for people to disclose personal information [Te Pas et al., 2020, Bhakta et al., 2014]. Our analysis of 42,332 patient feedback supports these findings, indicating that individuals are finding the new AI-enabled self-referral tool easy and convenient. This can be attributed to the tool’s flexibility, as individuals can refer to the service outside of the usual hours and take the necessary time to complete the referral process. Furthermore, the AI-enabled self-referral tool automatically opens on the service’s website, eliminating the need for patients to search for contact information or referral form. Another advantage is that individuals can more comfortably discuss their mental health issues without facing potential communication difficulties with a human. Addressing these barriers with an AI-enabled self-referral tool has likely lowered the threshold for seeking help, resulting in higher referral rates as indicated by our data.

Minority groups often face especially high barriers when seeking mental health treatment [Clement et al., 2015, Thornicroft et al., 2022], and our NLP analysis of patient feedback evaluated the mechanism of how the AI-enabled self-referral tool facilitated the increased referrals from minority groups. Interestingly, we found that minority groups mention different themes in their feedback, suggesting that the barriers they face and their needs are different to majority groups. We found that individuals from sexuality and gender minorities particularly valued the human-free nature of the self-referral tool, which enabled them to seek help without fear of judgment or discrimination. Ethnic minorities mainly mentioned how the referral process helped them to realise that they might be suffering from mental health problems or the need for treatment which may reflect the negative attitudes and lack of perceived need for treatment these individuals might face. Furthermore, minority groups expressed a greater need for specific treatments, highlighting the importance of addressing their unique needs and barriers. These final points indicate that while these initial results are reassuring, further work needs to be done to address all barriers experienced by minority groups when accessing mental health treatment.

In interpreting the results of the study, it is important to consider certain limitations. One potential issue to consider is the possibility of cultural differences between various NHS Talking Therapies services. It is possible that those NHS Talking Therapies services making use of the AI-enabled self-referral tool are inherently more open to digital health approaches and innovation than those that have not adopted such tools, which could influence the referral numbers. Nevertheless, implementation of the AI-enabled self-referral tool was not part of wider digital innovation programs for any of the services and the increase in referrals was tightly linked to the launch of the AI-enabled self-referral tool.

Despite this limitation, our study also has noteworthy strengths. As the data was collected as part of a standard national data reporting process that undergoes quality checks, there is no concern about missing data or that the data is reported in different ways. In addition, the data was collected under real-world conditions from multiple services, which provides a larger sample size and thereby improves the generalisability of the results. Furthermore, the staggered design of the self-referral tool implementation at different time points accounted for any natural changes in referral numbers. Our pre- versus post-period design and comparison to matched NHS Talking Therapies services further ensured that the observed results are not due to general external effects or other time-dependent factors or confounding factors.

The findings of this research demonstrate the potential of AI-enabled conversational tools to facilitate the self-referral process for mental health services and make it more accessible for different groups of people. This is particularly timely in light of the World Health Organisation’s Global strategy on digital health [World Health Organization, 2021a] and many healthcare providers around the world setting goals, such as the NHS Long-Term Plan in the UK [National Health Service, 2019], aiming to increase the number of people accessing mental health services in accordance with the UN 2030 Sustainable Development Goals (SDGs). Our results further support the notion that digital technologies can support reaching the SDGs [Lancet, 2021] and support more people in accessing the services they need. Utilizing an AI-enabled self-referral tool could reduce barriers to access, resulting in more referrals and improved treatment efficacy [Rollwage et al., 2022] and a decrease in the burden on individuals and society.

## Data Availability

Code and data in the present study are available upon reasonable request to the authors. The qualitative feedback data will not be available because the information could compromise participants' privacy/consent.

## 5 Acknowledgments

We thank Emili Ivanova for her contribution to the figures and illustrations.

## 7 Data and code availability

Code and data regarding the referral analysis will be made available upon submission acceptance. The qualitative feedback data will not be publicly available because the information could compromise participants’ privacy/consent.

