## Supplementary material for "Closing the accessibility gap to mental health treatment with a conversational AI-enabled self-referral tool"

#### Total number of self-referrals

To ensure that the observed increase in the total number of referrals in the manuscript was not due to other forms of referrals (e.g., GP referrals) increasing during the study period, we conducted a secondary analysis of only self-referrals. We repeated the analysis reported in the manuscript for self-referrals and used data from the same services during the same time period.

The implementation of the AI-enabled self-referral tool resulted in an 18% increase in the total number of self-referrals at the same time period as reported in the manuscript, whereas the other services had a 7% increase in self-referrals in the same time period (Figure 1).

The increase in total self-referral numbers for services using the AI-enabled self-referral tool was significantly higher than the increase for other similar services ($\chi^2$ (1) = 106.5, $p < 0.001$). This suggests that the self-referral tool facilitates an increase in self-referrals.

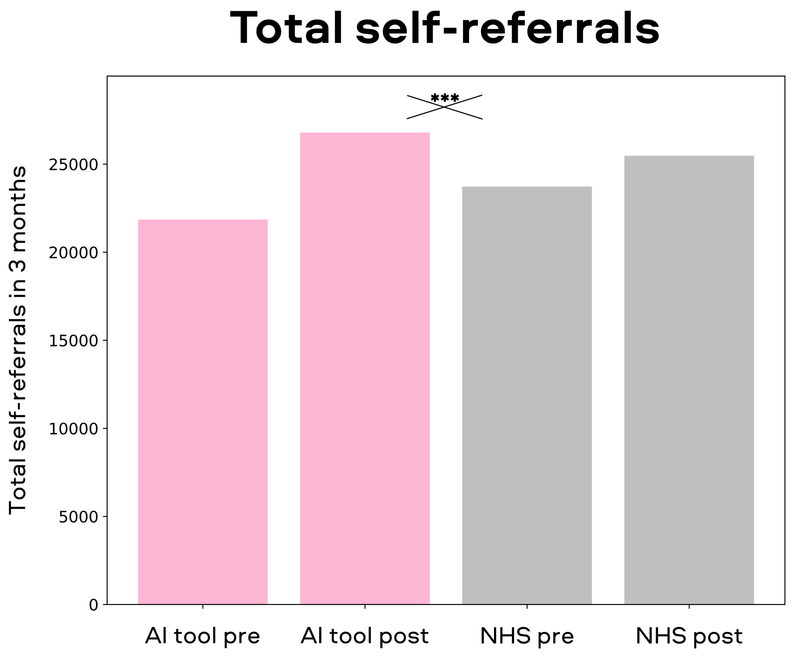


Supplementary Figure 1: The total number of self-referrals pre- and post-implementation of the AI-enabled self-referral tool for services that implemented the novel tool (pink) and other similar services that did not implement the tool (grey). There was a 18% increase in self-referrals for services that used the AI tool, whereas other services had a 7% increase in the same time period. The increase in self-referrals for services using the self-referral tool was significantly higher than the increase for other similar services.***$p < 0.001$

#### Total number of referrals split by demographic group

| | | **Total referrals pre AI tool** | **Total referrals post AI tool** |
|---|---|---|---|
| **Gender identity** | Female | 20735 | 24520 |
| | Male | 9775 | 11300 |
| | Non-binary | 100 | 335 |
| | Not answered/unknown | 70 | 70 |
| **Sexual orientation** | Heterosexual | 24560 | 27935 |
| | Bisexual | 1840 | 2395 |
| | Homosexual | 445 | 530 |
| | Not answered/unknown | 3880 | 5385 |
| **Ethnic group** | White | 26230 | 30065 |
| | Asian | 1000 | 1390 |
| | Mixed | 840 | 970 |
| | Black | 565 | 800 |
| | Other | 255 | 315 |
| | Not answered/unknown | 1635 | 2525 |

Supplementary Table 1: Total number of referrals pre-implementation of the AI-enabled self-referral tool and post-implementation of the tool. These numbers were used to calculate the percentage changes reported in the manuscript.

#### Number of referrals categorised by demographic group and feedback theme

| Theme | Number of referrals split by gender identity | | $\chi^2$ | Corrected p value |
|---|---|---|---|---|
| | Other | Females and Males | | |
| **Convenient** | 434 | 23948 | 7.79 | 0.142 |
| **Provided hope** | 84 | 5768 | 8.84 | 0.079 |
| **Self-realisation** | 65 | 3175 | 0.04 | 22.47 |
| **Human-free** | 113 | 3691 | 22.7 | 0.00005 |
| **Needed specific support** | 45 | 1624 | 4.8 | 0.77 |
| **Other neutral feedback** | 33 | 1248 | 2.46 | 3.148 |
| **Expected support sooner** | 20 | 828 | 0.58 | 12.02 |
| **Wanted urgent support** | 14 | 484 | 1.57 | 5.69 |
| **Other negative feedback** | 5 | 297 | 0.02 | 23.74 |

Supplementary Table 2: The number of referrals, categorised by gender identity and feedback theme, along with the corresponding Chi Square and corrected p values for comparing the gender minority group with females and males for each theme.

| Theme | Number of referrals split by sexual orientation | | $\chi^2$ | Corrected p value |
|---|---|---|---|---|
| | Bisexual | Heterosexual | | |
| **Convenient** | 1621 | 20110 | 0.72 | 10.68 |
| **Provided hope** | 306 | 4992 | 27.8 | $<0.00001$ |
| **Self-realisation** | 192 | 2691 | 3.49 | 1.671 |
| **Human-free** | 369 | 2980 | 62.4 | $<0.00001$ |
| **Needed specific support** | 162 | 1257 | 30.8 | $<0.00001$ |
| **Other neutral feedback** | 78 | 1021 | 0.28 | 16.08 |
| **Expected support sooner** | 40 | 679 | 3.90 | 1.307 |
| **Wanted urgent support** | 27 | 403 | 0.84 | 9.671 |
| **Other negative feedback** | 16 | 239 | 0.43 | 13.74 |

Supplementary Table 3: The number of referrals, categorised by sexual orientation and feedback theme, along with the corresponding Chi Square and corrected p values for comparing the bisexual individuals with heterosexual individuals for each theme.

| Theme | Number of referrals split by ethnic group | | $\chi^2$ | Corrected p value |
|---|---|---|---|---|
| | Asian and Black ethnic groups | White | | |
| **Convenient** | 1343 | 21905 | 24.2 | 0.00002 |
| **Provided hope** | 322 | 5253 | 2.74 | 2.64 |
| **Self-realisation** | 281 | 2783 | 46.6 | <0.00001 |
| **Human-free** | 215 | 3416 | 0.82 | 9.83 |
| **Needed specific support** | 133 | 1408 | 14.6 | 0.00361 |
| **Other neutral feedback** | 84 | 1106 | 1.12 | 7.83 |
| **Expected support sooner** | 67 | 709 | 7.02 | 0.217 |
| **Wanted urgent support** | 36 | 428 | 1.49 | 6.00 |
| **Other negative feedback** | 18 | 265 | 0.0 | 27.0 |

Supplementary Table 4: The number of referrals, categorised by ethnic group and feedback theme, along with the corresponding Chi Square and corrected p values for comparing the Asian and Black ethnic groups with the White group for each theme.

### Feedback comparison between all minority groups and majority group

To ensure that the feedback from all minority groups was consistent with the minority groups where we saw an increase in total referrals, we conducted a supplementary analysis that included all minority groups, not just those where we observed a significant increase in referrals compared to the majority group, as reported in the main manuscript. The findings from this supplementary analysis were consistent with those reported in the main manuscript.

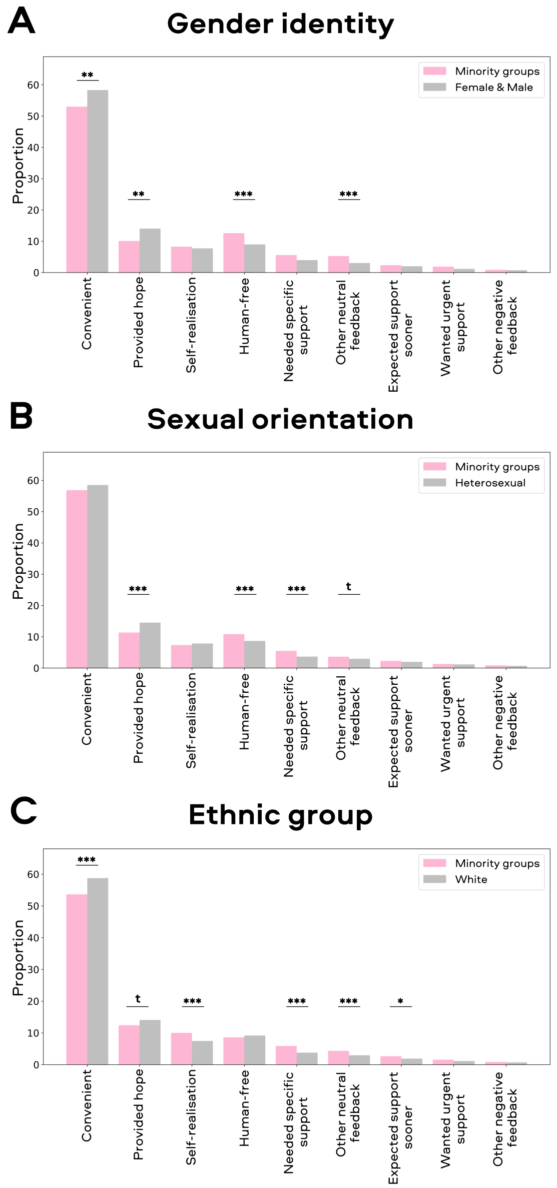


Supplementary Figure 2: Breakdown of feedback themes from minority (pink bars) and majority (grey bars) groups. A) Breakdown of feedback for gender minority groups compared to females and males. B) Breakdown of feedback for sexuality minority groups compared heterosexual individuals. C) Breakdown of feedback for ethnic minority groups compared White individuals. ***$p < 0.001$; ** $p < 0.01$; t $p < 0.1$

#### Access to clinical assessment

To ensure that the increase in the number of referrals did not result in longer wait times for clinical assessment and a reduction in the number of people accessing these assessments, we compared the percentage of people who were assessed during the study between the services that used the AI-enabled self-referral tool and services that did not use the tool. We conducted a two-way ANOVA to investigate the effects of the time period (i.e., pre- and post-implementation of the AI-enabled self-referral tool) and tool usage on the number of clinical assessments. The analysis showed a significant effect of the time period ($F(1, 13) = 4.67$, $p = 0.035$), indicating a reduction in the number of assessments over time. However, there was no significant effect of tool usage ($F(1, 13) = 0.06$, $p = 0.803$), and no significant interaction between time period and tool usage ($F(1, 13) = 0.09$, $p = 0.764$), suggesting that the AI-enabled self-referral tool did not have a negative impact on the number of assessments that were conducted.

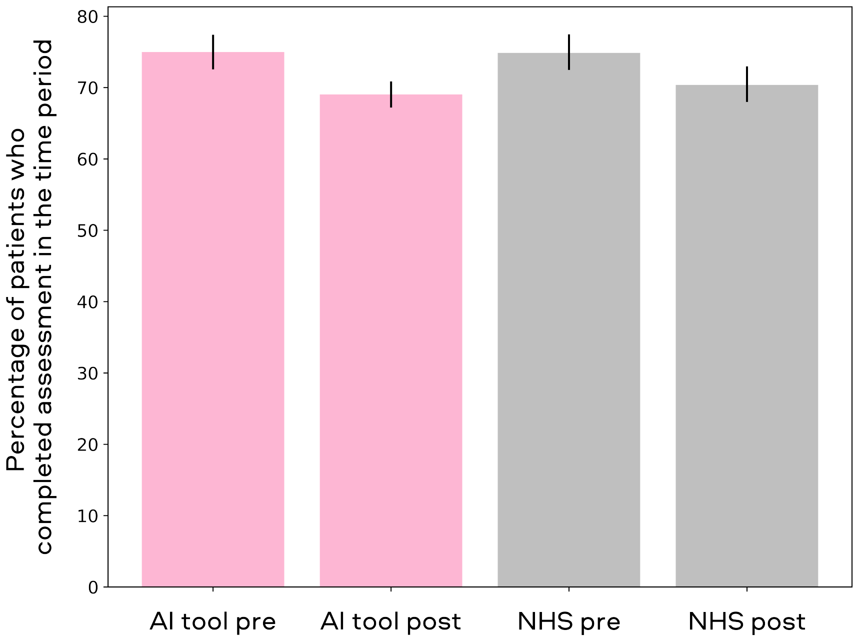


Supplementary Figure 3: The percentage of individuals who went through a clinical assessment in the study time period for services that implemented the novel AI-enabled self-referral tool (pink) and other similar services that did not implement the tool (grey).
